# Trends in syphilis prevalence by race and ethnicity among people who are pregnant in the United States 2016–2023

**DOI:** 10.1101/2024.07.25.24310963

**Authors:** Yizhi Liang, Nicolas A Menzies, Minttu M Rönn

## Abstract

**Background:** This study aimed to estimate syphilis prevalence among people who are pregnant with live births by race and ethnicity 2016–2023.

**Methods:** We analyzed data on syphilis infection from U.S. birth certificates using a Bayesian mathematical model, adjusting for test sensitivity, specificity, and screening coverage. We calibrated the model under two scenarios: (1) assuming screening coverage is as estimated in Medicaid claims data and (2) assuming higher screening coverage than observed in Medicaid claims data. We compared the estimates to stillbirths attributable to syphilis reported through routine surveillance. We examined racial and ethnic disparities using the index of disparity.

**Results:** In Scenario 1, syphilis prevalence among people who are pregnant with live births increased from 101.1 (95% uncertainty interval [UI]: 87.5–120.5) per 100,000 live births in 2016 to 533.4 (95% UI: 496.6–581.0) per 100,000 live births in 202. In Scenario 2, prevalence increased from 73.9 (95% UI: 51.2–130.0) per 100,000 live births to 378.1 (95% UI: 295.5–592.0) per 100,000 live births over the same period. With rising prevalence, relative racial and ethnic disparities narrowed over time. Prevalence was estimated to be higher among women with stillbirths compared to women with live births.

**Conclusions:** In the United States, improved estimates of screening coverage are needed to understand the gaps in congenital syphilis prevention and to inform estimates of syphilis prevalence among pregnant persons.

**One sentence summary line:** In the United States, we estimated an increasing syphilis prevalence among people who are pregnant, reaching 533 per 100,000 live births in 2023, with notable racial and ethnic disparities.

## Introduction

Syphilis is a sexually transmitted infection (STI) caused by the bacterium *Treponema pallidum*.^1^ After reaching a low of 2.1 diagnoses per 100,000 persons in 2001, the syphilis diagnosis rate in the United States has been on the rise.^2^ In 2023, the total syphilis diagnosis rate in the general population was 2.3 times that of 2019.^3^ Syphilis can be transmitted vertically during pregnancy, resulting in congenital syphilis, which can lead to stillbirth, neonatal death, and other adverse birth outcomes.^4^ The rate of congenital syphilis has also been increasing, and in 2023, it was 6.5 times that of 2016.^3^ The rate of syphilis in people who are pregnant has increased in parallel, as recorded in the National Vital Statistics System’s natality data files, which compile information from birth certificates for all births occurring in the United States. Between 2016 and 2023, syphilis detections increased from 87.1 to 321.8 per 100,000 live births in natality data files. An increase was observed across all ages and racial and ethnic groups, with the largest increase in American Indian and Alaska Native people who are pregnant.^5^ Despite recommendations indicating a minimum of one syphilis test during pregnancy, estimates reported in Medicaid claims data indicate that screening coverage remains below recommendations,^6^ and no testing or untimely testing was indicated in 40% of congenital syphilis cases in 2022.^7^ It is likely that syphilis detected in birth certificate data underestimates the prevalence of syphilis in people who are pregnant. Birth certificate data do not record information on syphilis testing, but mathematical modeling can be used to estimate the prevalence of infection if we have external information about testing coverage in a given population. For example, a mathematical modeling analysis estimated that syphilis incidence was 50% higher than the number of reported diagnoses in the US.^8^

In this study, we aimed to estimate syphilis prevalence among people who are pregnant and who delivered live births in the United States from 2016 to 2023. We leveraged information on maternal race and ethnicity and detected syphilis infections in birth certificate data, together with information on syphilis screening coverage and test sensitivity and specificity. We evaluated the trends in racial and ethnic disparities over time. Quantifying the total prevalence of infection can aid syphilis prevention efforts and improve health. Prevalence time trends can also improve understanding of the magnitude and speed of syphilis re-emergence in different populations.

## Methods

### Birth certificate data

We used data from birth certificates of the United States, as maintained within the National Vital Statistics System by the National Center for Health Statistics (NCHS) and accessible from the National Bureau of Economic Research (https://www.nber.org/research/data/vital-statistics-natality-birth-data). These records include demographic and health-related microdata for all live births occurring within a calendar year, as mandated by the birth registration requirements across the 50 states, New York City, and the District of Columbia^9,10^. Non-single delivery status is not identifiable in the birth certificates,^9^ so we assumed that each record represents an individual birth. Revised 2003 U.S. Standard Certificate of Live Birth was introduced to improve data quality,^11^ and by January 1, 2014, 96.2% of all births to U.S. residents were documented utilizing the 2003 standard.^12^ We restricted our analysis to 2016–2023 to follow the years covered in the National Center for Health Statistics Brief on trends in maternal syphilis rates during pregnancy.^5^

We defined seven racial and ethnic categories: non-Hispanic American Indian/Alaska Native (AIAN), non-Hispanic Asian (Asian), non-Hispanic Black/African American (Black), Hispanic/Latino (Hispanic), non-Hispanic Multiracial (Multiracial), non-Hispanic Native Hawaiian/Other Pacific Islander (NHPI), and non-Hispanic White (White).

Syphilis infection status reported in the birth certificates is extracted from the medical records utilizing the Facility Worksheets,^13,14^ following the protocols outlined in the “Guide to Completing the Facility Worksheets for the Certificate of Live Birth and Report of Fetal Death (2003 Revision).”^15^ The identification of syphilis infection (also referred to as lues) was determined by: i) a positive test for *Treponema pallidum* presenting at the start of pregnancy or confirmed diagnosis during pregnancy with or without documentation of treatment; ii) the presence of documented treatment for syphilis during pregnancy was deemed sufficient in the absence of a definitive diagnosis within the accessible records.^15^

### Statistical modeling

We estimated syphilis prevalence among people who are pregnant with live births stratified by year and race and ethnicity using a Bayesian mathematical modeling approach. The model is anchored on the number of detected syphilis infections observed in birth certificate data in each race and ethnicity and year stratum. We modeled the probability of observing a positive syphilis infection among people who are pregnant θ_*yr*_ as:

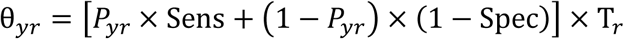

Where *P*_*yr*_ is syphilis prevalence among people who are pregnant stratified by race and ethnicity and year, Sens and Spec are the sensitivity and specificity of syphilis diagnostic testing; both assumed constant by year and population. T_*r*_ is the proportion of people who are pregnant screened for syphilis during their pregnancy stratified by race and ethnicity but assumed constant over 2016-2022.

Parameters and their prior distributions are presented in Table 1Syphilis testing follows an algorithm that can include multiple tests. To examine test sensitivity and specificity, we conducted a random-effect meta-analysis to pool sensitivity and specificity on estimates reported in a systematic review (Appendix Figure S1 and Figure S2).^16^ The pooled estimates indicated high sensitivity (0.98; 95% uncertainty range: 0.97–0.99) and specificity (0.99; 95% uncertainty range: 0.98–0.99), used in the analysis.

**Table 1.**
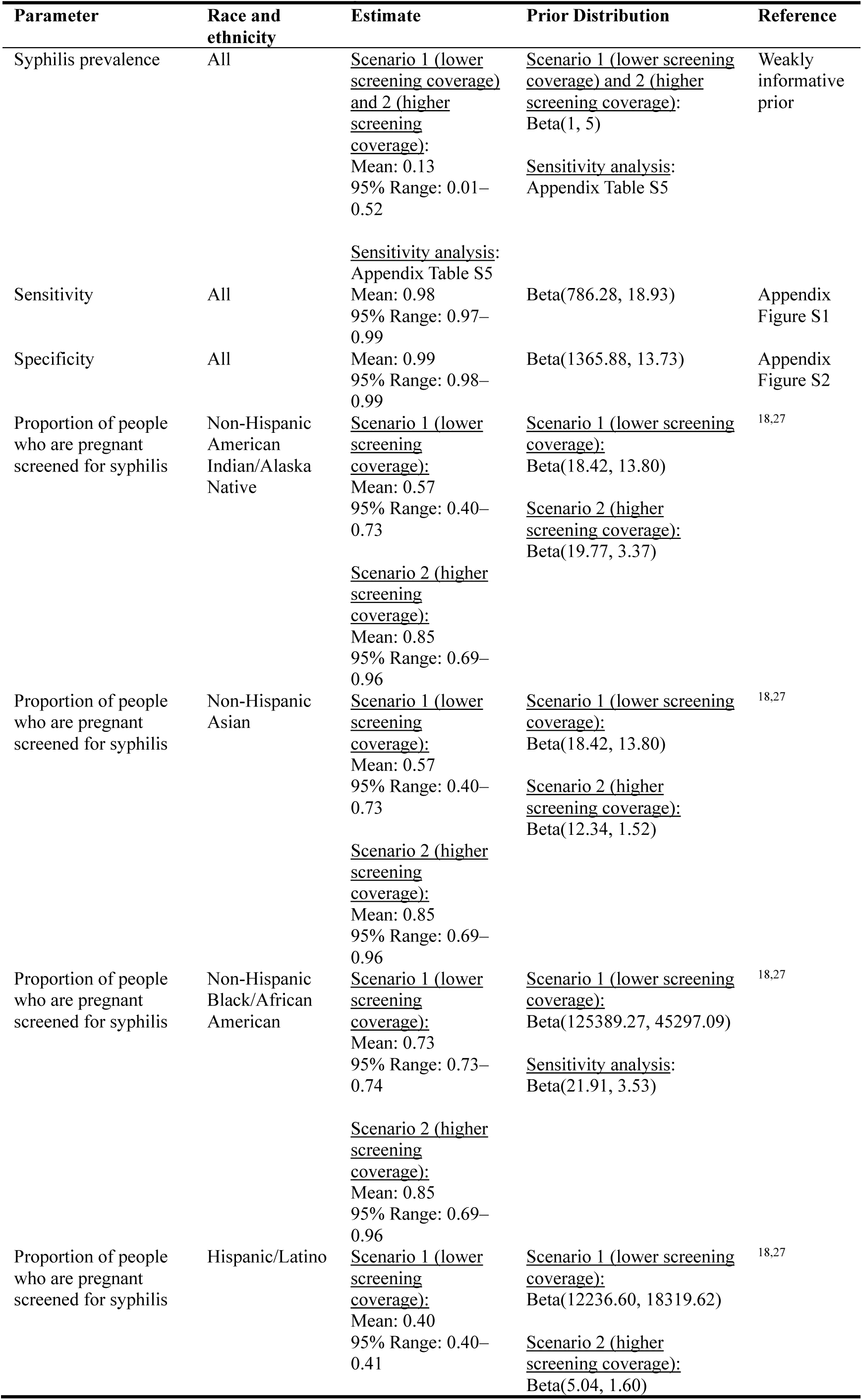
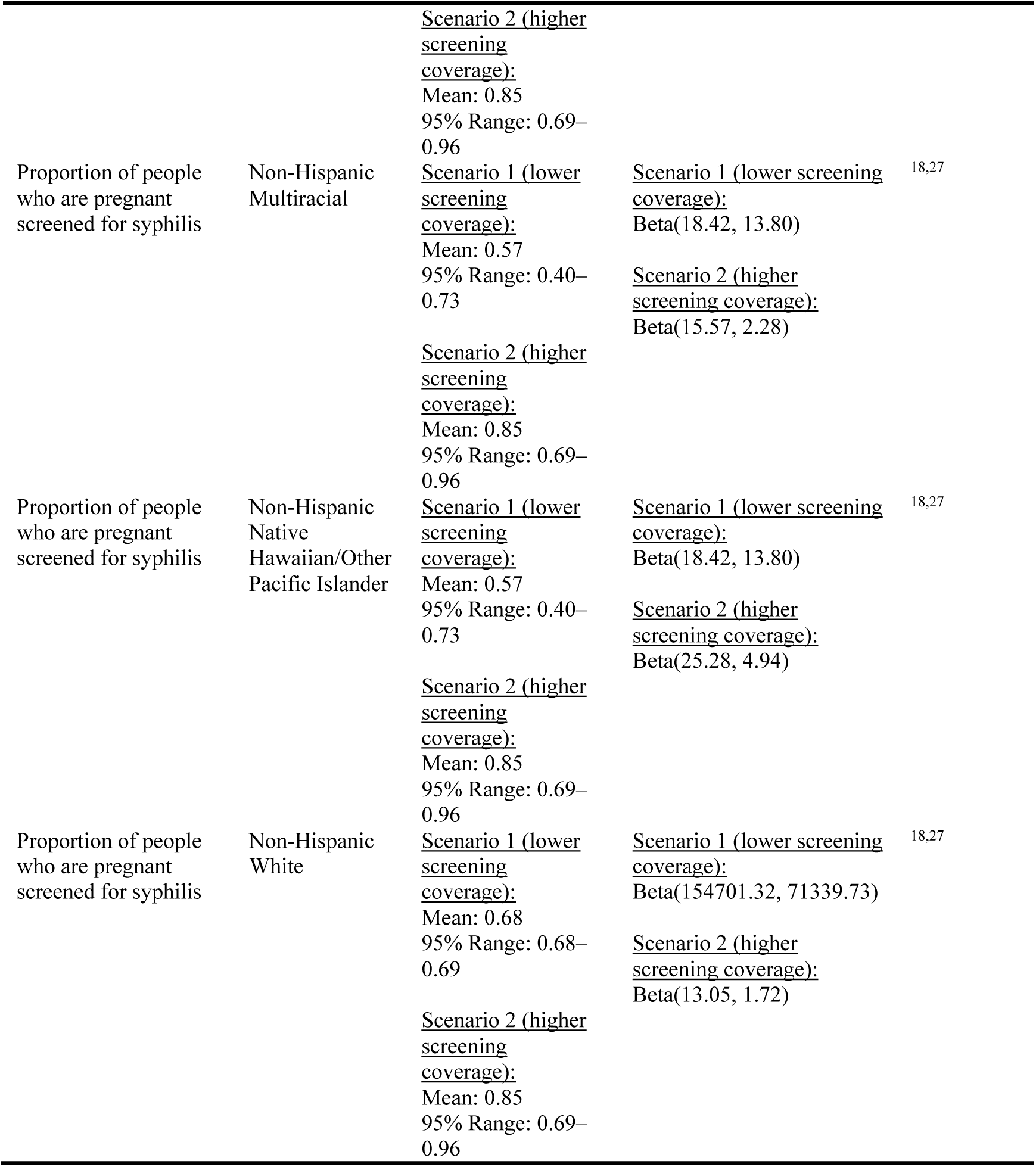
Parameters used in the analysis for 2016–2023.

There is limited information on syphilis screening coverage among people who are pregnant by race and ethnicity. Additionally, there are likely racial and ethnic disparities in syphilis screening, with a higher proportion of congenital syphilis cases occurring in AIAN and NHPI having no or untimely testing reported.^17^ Screening coverage is a key parameter in estimating the prevalence of a condition, and we calibrated the model under two scenarios. In the first scenario, referred to as “lower screening coverage” (Scenario 1), we parameterized the prior distribution of screening probability based on a study of syphilis screening coverage among Medicaid-insured pregnant people.^18^ The study reported screening coverage for White, Black, and Hispanic women. To determine prior distribution for AIAN, Asian, Multiracial, and NHPI women, we used the lower and higher 95% confidence intervals for Black and Hispanic women, who had the highest and lowest screening coverage, respectively.

Medicaid claims data may underestimate syphilis screening coverage in people who are pregnant if their screening was not billed to Medicaid or not billed in a format that itemizes services provided.^6,19^ In the second scenario, referred to as “higher screening coverage” (Scenario 2), we allowed for more uncertainty and higher screening coverage using a wider uncertainty range in the prior distribution: the 2.5th percentile of the prior distribution reflected reported Medicaid screening coverage and the 97.5th percentile represented the proportion of people who are pregnant receiving any prenatal care from birth certificates (Table 1). The upper level assumes that all women who received prenatal care received syphilis screening.

We calibrated the model to the observed syphilis infections in birth certificate data using binomial likelihood:

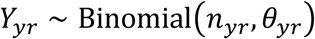

Where *Y*_*yr*_ is the number of detected syphilis in birth certificate data by year and race and ethnicity (Appendix Table S1). *n*_*yr*_ represents the number of women with live births by year and race and ethnicity (Numbers reported in Appendix Table S2).

The model was developed in R (version 4.3.2) and Stan (version 2.26.1) with “rstan”^20^, “tidybayes”^21^, and “ggplot2”^22^ packages. Calibration was performed via Markov Chain Monte Carlo sampling using the No-U-Turn Sampler (NUTS) ^23^. We specified 12,000 iterations per chain across four independent chains, with the first 8,000 designated as burn-in. Convergence of the model was assessed using the potential scale reduction factor (r-hat), where 1.1 was set as the indication of convergence.^24^ The analytic code is available at: [https://github.com/Yizhi-Liang/Trends-Syph].

### Comparison against external measures of burden

We compared the modeled prevalence estimates and detected syphilis infections in the birth certificate data against syphilis diagnoses among women of reproductive age (15– 44 years)^25^ and syphilis diagnoses among people who are pregnant^26^ (Appendix Table S3 and Table S4). Birth certificate data allows estimation of syphilis prevalence among people who are pregnant and delivered live births. Given that vertical transmission of syphilis increases the risk of stillbirth, the syphilis burden may be higher among pregnancies that did not result in live births. To further evaluate this, we obtained stillbirths attributable to congenital syphilis^26^ and the total number of stillbirths reported as part of fetal death data from the NCHS.^27^ We calculated the rate of syphilis attributable to stillbirths and compared this to the prevalence estimate among live births. This provides a crude estimate of the syphilis burden not measured in the analyses.

### Analysis

Using 16,000 posterior samples, we calculated the mean and 95% uncertainty intervals (UIs) to estimate syphilis prevalence by race and ethnicity and year. We examined temporal trends in syphilis prevalence by comparing changes in estimated syphilis prevalence over time, with 2016 used as the baseline. To evaluate racial and ethnic disparities, prevalence ratios were computed with White women, who had the largest number of live births, used as the reference population. We computed the Index of Disparity to quantify the variance in the estimated syphilis prevalence across races/ethnicity populations relative to the population mean, serving as a comprehensive measure of relative disparities:^28^

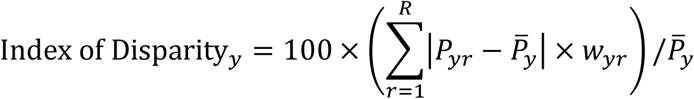

Where *P*_y*r*_ is the estimated syphilis prevalence for each race and ethnicity population in a year, *P̅_y_* is the average of the estimated syphilis prevalence for the whole population in the same analytical year, and *w*_*yr*_ is the population share for each race and ethnicity in each year.

### Sensitivity analysis

We tested alternative prior distributions for the syphilis prevalence by year and race/ethnicity to examine the impact of the shape of the weakly informative prior distribution on the posterior distribution (Appendix Table S5).

## Results

We analyzed syphilis prevalence estimates in relation to detected cases recorded on birth certificates, diagnoses reported among pregnant individuals, and those reported among women of reproductive age (15-49 years). In the lower screening coverage scenario, the estimated prevalence of syphilis was notably higher compared to other measures, while the prevalence in the higher screening coverage scenario aligned closely with the estimates from birth certificate data. When examining the reported diagnosis rates among pregnant women, the lower screening coverage scenario showed a prevalence-to-diagnosis ratio of 1.89 for 2023, while the higher screening coverage scenario had a ratio of 1.34 (Figure 1). Over the years, the discrepancy between reported syphilis diagnoses and cases detected on birth certificates diminished, converging in 2021.

**Figure 1.**
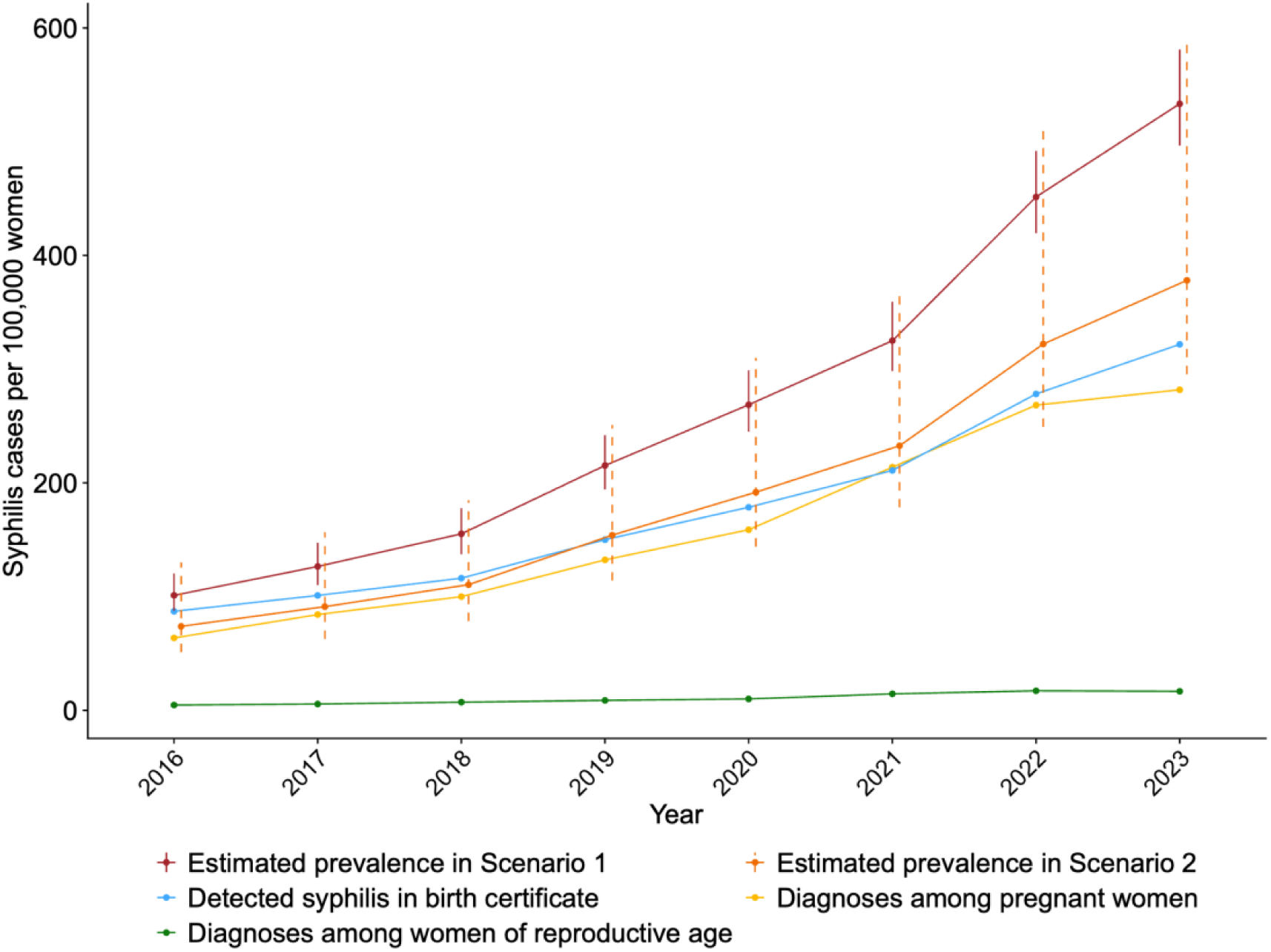
Comparison of the estimated syphilis prevalence per 100,000 live births, detected syphilis in birth certificate data per 100,000 live births, syphilis diagnoses among people who are pregnant per 100,000 women, and syphilis diagnoses among women of reproductive age per 100,000 women for 2016–2023. Footnote: In Scenario 1, we assume a lower screening coverage distribution among Medicare beneficiaries of each race/ethnicity. In Scenario 2, we assume a higher screening coverage distribution, ranging from the coverage reported by Medicare to the proportion of pregnant women receiving at least one prenatal care.

We observed an increase in syphilis cases across all racial and ethnic populations from 2016 to 2023 in both scenarios (Figure 2). When comparing these prevalence estimates to the syphilis cases detected in birth certificates, the estimates from the higher screening coverage scenario indicated a burden of syphilis that closely resembled what had been observed. However, in certain cases, the prevalence estimates were lower than those detected in birth certificates. For instance, the higher screening coverage scenario suggested that the syphilis prevalence among pregnant Asian individuals was less than detected in the birth certificate data. This discrepancy implied the possibility of a number of false positive syphilis cases in the birth certificate records.

**Figure 2.**
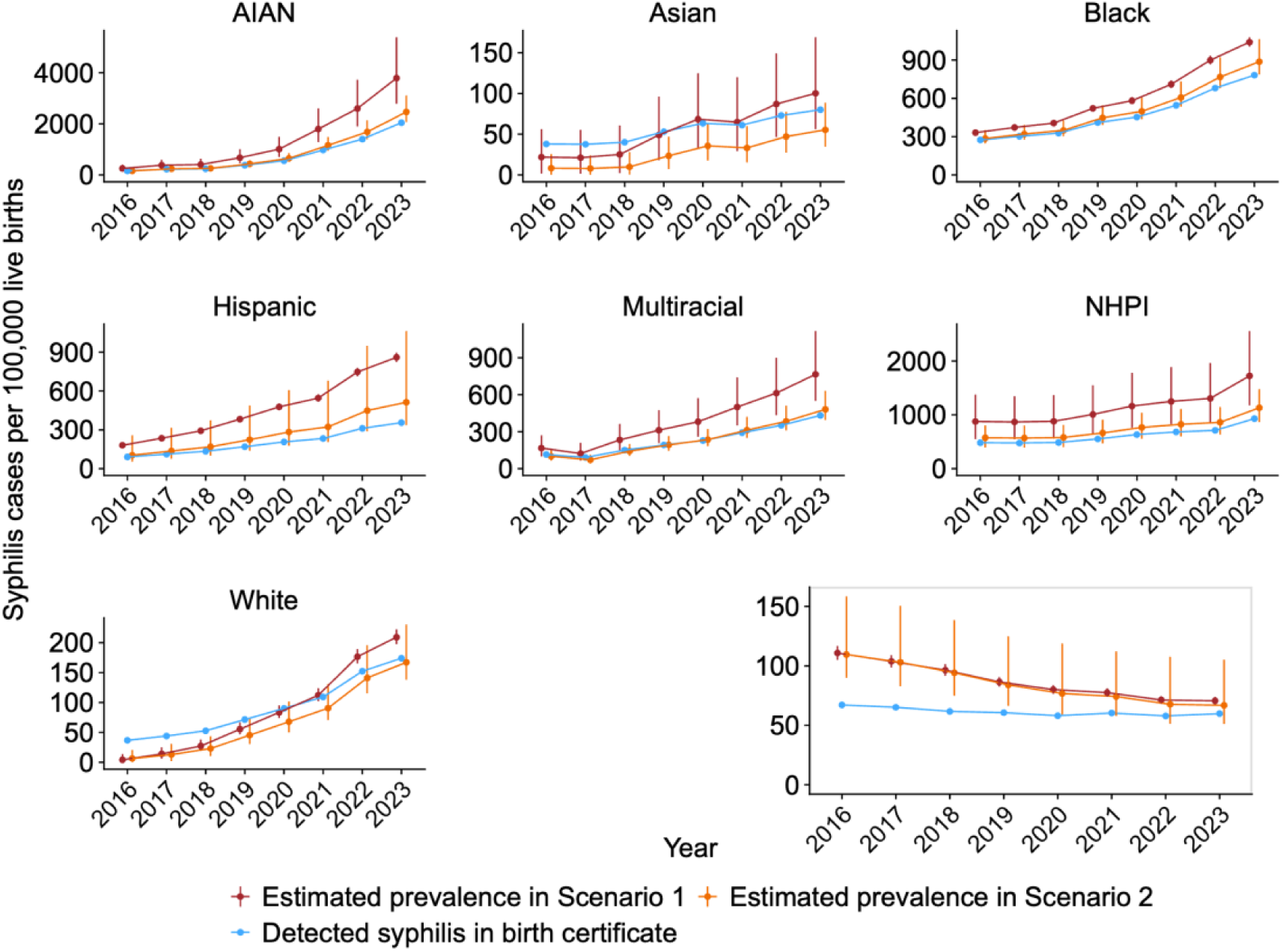
Comparison between the estimated syphilis prevalence per 100,000 live births and the detected syphilis in birth certificate data per 100,000 live births for 2016–2023. The y-axis varies by subplot. The Index of Disparity by year across race and ethnicity is shown in the right-bottom corner. Footnote: In Scenario 1, we assume a lower screening coverage distribution among Medicare beneficiaries of each race/ethnicity. In Scenario 2, we assume a higher screening coverage distribution, ranging from the coverage reported by Medicare to the proportion of pregnant women receiving at least one prenatal care. Race and ethnicity categories are non-Hispanic American Indian/Alaska Native (AIAN), non-Hispanic Asian (Asian), non-Hispanic Black/African American (Black), Hispanic/Latino (Hispanic), non-Hispanic Multiracial (Multiracial), non-Hispanic Native Hawaiian/Other Pacific Islander (NHPI), and non-Hispanic White (White).

The syphilis prevalence ratio comparing 2023 with 2016 was 5.2 in both scenarios: in 2016, the prevalence was 101.1 (95% UI: 87.5–120.5) per 100,000 live births, while in 2023, it reached 533.4 (95% UI: 496.6–581.0) per 100,000 live births in the lower screening coverage scenario; they were 73.9 (95% UI: 51.2–130.0) in 2016 and 378.1 (95% UI: 295.5–592.0) in 2023 per 100,000 live births in the higher screening coverage scenario. In the lower screening coverage scenario, the highest prevalence in 2016 was among NHPI women at 875.9 (95% UI: 547.6–1376.1) per 100,000 live births, while Asian women had the lowest at 22.0 (95% UI: 1.9–56.1) per 100,000 live births. This rank remained the same in the higher screening coverage scenario, where the prevalence for NHPI women was 574.0 (95% UI: 394.1–802.8), and for Asian women was 8.3 (95% UI: 0.3–25.7). In 2022, under both scenarios, AIAN women were estimated to have the highest prevalence (3790.5 per 100,000 live births, 95% UI: 2788.2–5384.5 in Scenario 1; 2466.2 per 100,000 live births, 95% UI: 2075.4–3111.5 in Scenario 2), followed by NHPI women (1722.2 per 100,000 live births, 95% UI: 1174.5–2557.4 in Scenario 1; 1130.8 per 100,000 live births, 95% UI: 865.2–1474.9 in Scenario 2) and Black women (1040.0 per 100,000 live births, 95% UI: 1003.5–1077.5 in Scenario 1; 886.9 per 100,000 live births, 95% UI: 786.5–1063.6 in Scenario 2). Asian women were estimated to have the lowest prevalence (100.2 per 100,000 live births, 95% UI: 56.2–168.9 in Scenario 1; 55.4 per 100,000 live births, 95% UI: 34.8–88.6 in Scenario 2).

All racial and ethnic populations, excluding Asian women, were estimated to have a higher syphilis prevalence compared to White women between 2016 and 2023. The absolute differences between populations widened, while the prevalence ratio decreased over time in all racial and ethnic groups. This pattern emerged due to the increase in syphilis prevalence estimated for the White population (Appendix Figure S3 and Figure S4). There was a decreasing trend in the disparity of syphilis prevalence across racial and ethnic groups in both scenarios and in syphilis detections observed in birth certificate data, while the tendency for the detected syphilis cases was not as obvious as those in the estimated prevalence, as measured by the Index of Disparity (Figure 2). In 2016, the index was 110.8, 109.6, and 67.1 in estimated prevalence in the lower screening coverage scenario, the higher screening coverage scenario, and detected syphilis cases in birth certificates, respectively. The corresponding values in 2023 were 70.7, 66.8, and 59.8. The Index of Disparity decreased due to the rising prevalence across all racial and ethnic populations.

In a sensitivity analysis, prevalence estimates were robust under different weakly informative prior distributions, maintaining stability even when the prior distributions’ central values and the 95% uncertainty ranges were increased (Appendix Figure S5)

When we compared our estimates of syphilis prevalence among live births to the prevalence among stillbirths based on stillbirths reported in surveillance data, the estimated syphilis prevalence among live births was consistently lower than the proxy estimates for stillbirths, with the discrepancy widening over the years (Figure 3). In 2023, the syphilis prevalence estimated among stillbirths based on surveillance data was 1336.0 per 100,000 births, which was 2.5 (95% UI: 2.3–2.7) times the estimated syphilis prevalence among those with live births in lower screening coverage scenario, and 3.5 (95% UI: 2.3–4.5) times that of the prevalence in the higher screening coverage scenario.

**Figure 3.**
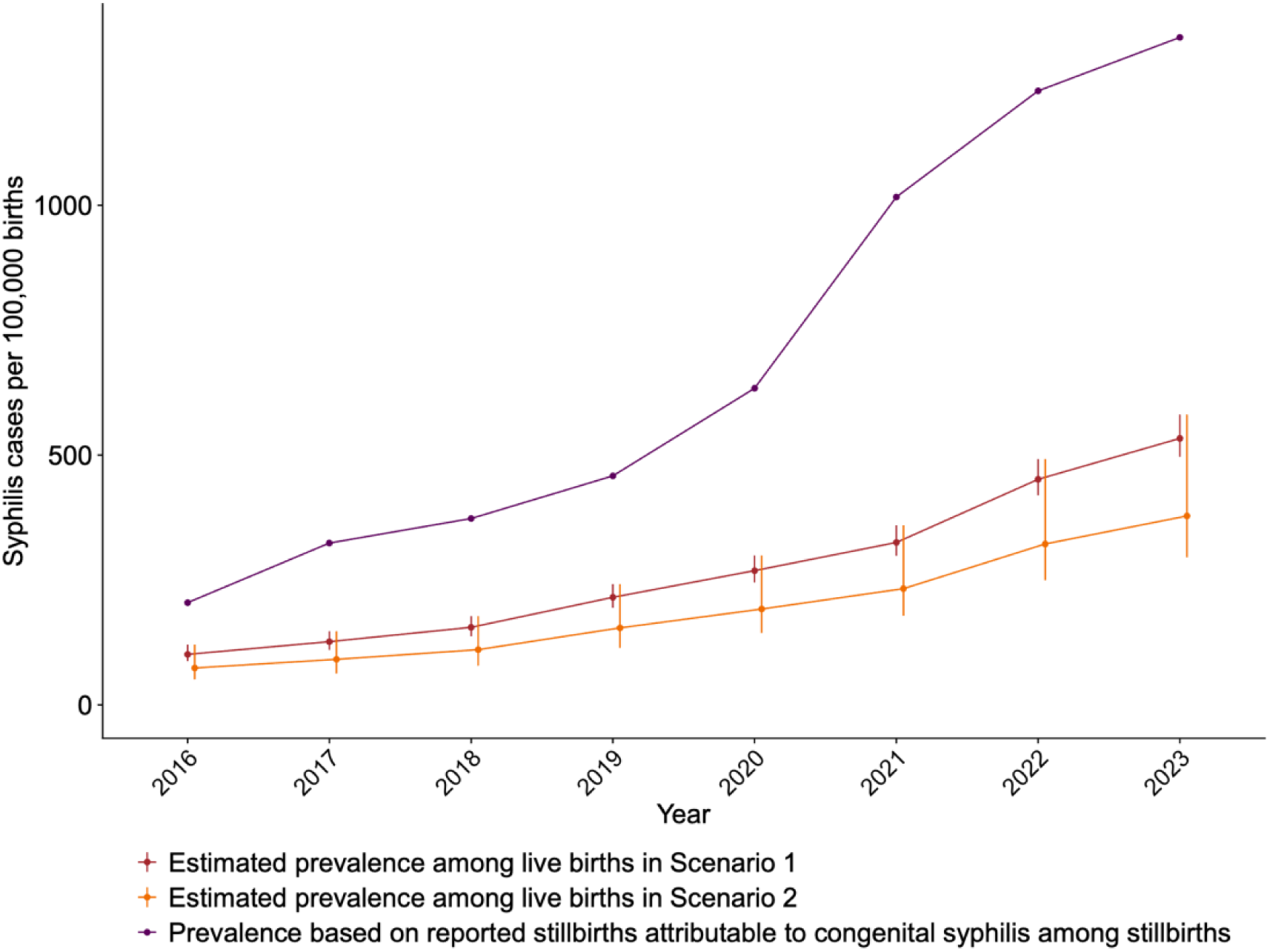
Comparison between the estimated syphilis prevalence per 100,000 live births and the prevalence based on reported stillbirths attributable to congenital syphilis among stillbirths per 100,000 stillbirths. Footnote: In Scenario 1, we assume a lower screening coverage distribution among Medicare beneficiaries of each race/ethnicity. In Scenario 2, we assume a higher screening coverage distribution, ranging from the coverage reported by Medicare to the proportion of pregnant women receiving at least one prenatal care.

## Discussion

We estimated an upward trend in syphilis prevalence associated with persistent racial and ethnic disparities between 2016 and 2023. Racial and ethnic disparities were predicted to increase over time on an absolute scale but diminish in relative terms, driven by increasing burden in all racial and ethnic groups. In addition, we estimated that syphilis prevalence in women who experience stillbirth is at least twice our estimated syphilis prevalence among women with live births.

The disparities observed are rooted in systemic racism and are perpetuated by socioeconomic factors such as poverty, inequalities in access to quality healthcare, and broader social determinants of health.^2^ In cases of congenital syphilis, the absence or delay of syphilis screening during pregnancy has been identified as a critical factor, particularly pronounced among AIAN and NHPI women.^17^ Improved estimates of screening coverage are needed to understand the gaps in congenital syphilis prevention. They are needed to inform the estimation of the underlying prevalence of syphilis in people who are pregnant. The Healthy People 2030 project has documented a decline in the proportion of people who are pregnant receiving early and adequate prenatal care across all groups, with NHPI and AIAN women experiencing the lowest level of adequate care in 2018–2022.^29^ Studies on racial and ethnic disparities in syphilis among people who are pregnant have focused on White, Black, and Hispanic populations. There remain data gaps for the smaller racial and ethnic populations, such as NHPI and AIAN, who are disproportionately affected by the poor quality of prenatal care and higher burden of syphilis.^3,18,30^

US Preventive Services Task Force recommends early syphilis screening for all people who are pregnant, with a further recommendation for an additional test during the third trimester for those at elevated risk.^31^ The American Academy of Pediatrics and the American College of Obstetricians and Gynecologists have also updated their guidelines, advocating for three syphilis screens (first trimester, third trimester, and at delivery) as part of routine prenatal care.^32^

The contribution of syphilis to stillbirths remains understudied. The identification of stillbirths attributable to congenital syphilis presents a challenge; different infectious diseases during pregnancy increase the risk of stillbirth, and there is inadequate adherence to syphilis screening at the time of stillbirth.^33^ Consequently, syphilis-attributable stillbirths reported to congenital syphilis surveillance may present an underestimate, and the discrepancy between syphilis prevalence in live births and stillbirths may be higher than estimated.

Our study leverages a comprehensive dataset representing almost all live births in the United States for 2016–2023. This allowed us to estimate prevalence for the smaller racial and ethnic populations, such as AIAN and NHPI, who experience disproportionate burden but are often not represented in analyses. Our findings accounted for imperfect test sensitivity and specificity. The estimates were broadly aligned and showed similar trends with surveillance data despite uncertainty around testing coverage by time and in different populations. However, our analysis was confined to pregnancies with live births, and it does not include the burden among stillbirths. Our analysis is at the national level. There is geographic variation in syphilis burden, and there may be variation in prenatal syphilis screening practices, which was not accounted for in this study.

This study provides evidence of the increasing syphilis burden among people who are pregnant with live births, demonstrating the increasing syphilis burden in all racial and ethnic populations in the United States and the presence of racial and ethnic disparities. Addressing these disparities is needed to improve inequalities in birth outcomes.

## Data Availability

All data produced are available online at https://github.com/Yizhi-Liang/Trends-Syph.

## Notes

Disclosures: No conflict to declare

### Competing Interest Statement

The authors have declared no competing interest.

### Funding Statement

This study did not receive any funding.

### Author Declarations

The study used only openly available data from published literature cited and databases that were originally located at the National Center for Health Statistics (https://www.cdc.gov/nchs/data_access/vitalstatsonline.htm), Sexually Transmitted Infections Surveillance 2023 (https://www.cdc.gov/sti-statistics/annual/index.html), and NCHHSTP AtlasPlus (https://www.cdc.gov/nchhstp/about/atlasplus.html?CDC_AAref_Val=https://www.cdc.gov/nchhstp/atlas/index.htm).

### Summary of Updates

We updated the analysis to cover the years from 2016 to 2023. Additionally, we calibrated the model under two key scenarios: (1) incorporating screening coverage as estimated in Medicaid claims data and (2) assuming a higher screening coverage than what was previously observed in those claims. All plots have been adjusted accordingly, and the supplemental files have been updated to reflect the changes in the analyzed year and scenarios.

