## Appendix for "Trends in syphilis prevalence by race and ethnicity among people who are pregnant in the United States 2016–2023"

**Meta-analysis of sensitivity and specificity of treponemal-specific tests for the diagnosis of syphilis among people who are pregnant.**

Park el al.[1] reported the performance of treponemal immunoassays for syphilis screening and summarized the sensitivity and specificity of these tests by syphilis stages. Building on their work, we conducted a random-effect meta-analysis using the "meta" package and utilized the generalized linear mixed model with the logit transformation to compute the overall sensitivity and specificity of these tests.[2] The values with the uncertainty range for the prior distributions for the sensitivity and specificity of the syphilis test received by people who are pregnant in the US were obtained and presented in Table 1 in the manuscript. The analytic code was shown in the "1_sens_spec.R" file under the "code/1_parameteriazation" folder via the GitHub repository mentioned in the manuscript.

### Figure S1. Forest plot for the sensitivity of syphilis test.

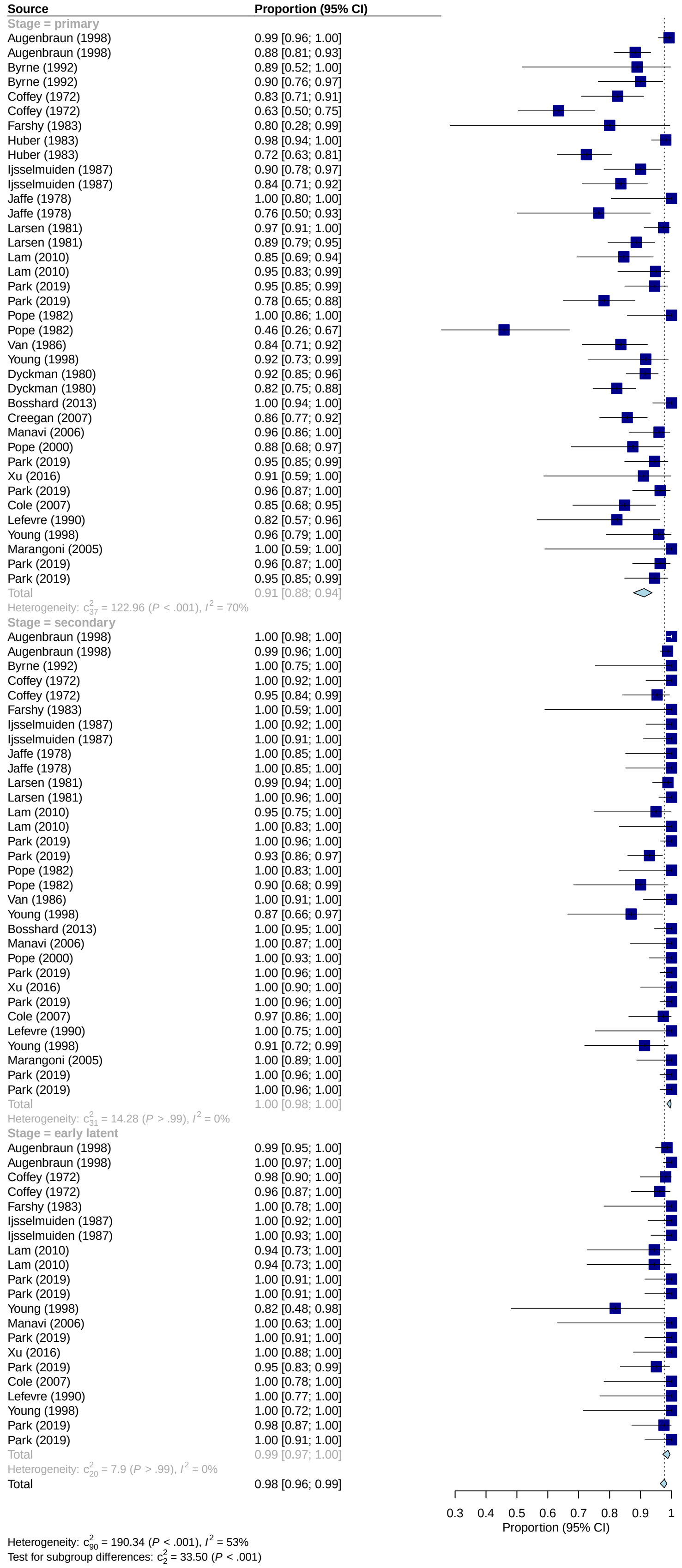

**Figure S2. Forest plot for the specificity of syphilis test.**

| Source | Proportion (95% CI) |
|---|---|
| **Type = manual treponemal** | |
| Byrne (1992) | 0.92 [0.84; 0.96] |
| Coffey (1972) | 0.99 [0.98; 1.00] |
| Farshy (1983) | 0.95 [0.92; 0.97] |
| Ijsselmuiden (1987) | 1.00 [0.99; 1.00] |
| Ijsselmuiden (1987) | 1.00 [0.99; 1.00] |
| Jaffe (1978) | 0.99 [0.98; 0.99] |
| Jaffe (1978) | 0.99 [0.98; 1.00] |
| Larsen (1981) | 0.98 [0.95; 0.99] |
| Larsen (1981) | 0.99 [0.97; 1.00] |
| Larsen (1981) | 0.95 [0.93; 0.97] |
| Larsen (1981) | 1.00 [0.99; 1.00] |
| Lan (2010) | 1.00 [0.92; 1.00] |
| Lan (2010) | 0.91 [0.78; 0.97] |
| Moyer (1984) | 0.96 [0.94; 0.98] |
| Park (2019) | 1.00 [0.99; 1.00] |
| Park (2019) | 0.98 [0.96; 0.99] |
| Pope (1982) | 0.99 [0.96; 1.00] |
| Pope (1982) | 0.99 [0.96; 1.00] |
| Romanowski (1987) | 0.87 [0.76; 0.94] |
| Van (1986) | 1.00 [0.99; 1.00] |
| Young (1998) | 1.00 [1.00; 1.00] |
| Bosshard (2013) | 0.99 [0.95; 1.00] |
| Cole (2007) | 0.99 [0.97; 1.00] |
| Liu (2014) | 1.00 [0.98; 1.00] |
| Pope (2000) | 0.95 [0.91; 0.98] |
| Wellinghausen (2011) | 1.00 [0.99; 1.00] |
| Total | 0.99 [0.98; 0.99] |
| Heterogeneity: $c^2_{25}$ = 142.23 ($P$ < .001), $I^2$ = 82% | |
| **Type = treponemal immunoassays** | |
| Park (2019) | 0.96 [0.93; 0.97] |
| Liu (2014) | 0.91 [0.86; 0.94] |
| Marangoni (2009) | 0.98 [0.98; 0.99] |
| Saral (2012) | 1.00 [1.00; 1.00] |
| Wellinghausen (2011) | 1.00 [0.99; 1.00] |
| Xia (2018) | 0.98 [0.97; 0.99] |
| Xu (2016) | 1.00 [1.00; 1.00] |
| Park (2019) | 0.97 [0.95; 0.98] |
| Cole (2007) | 1.00 [0.99; 1.00] |
| Silletti (1995) | 0.98 [0.96; 0.99] |
| Xia (2018) | 0.98 [0.97; 0.99] |
| Marangoni (2005) | 1.00 [1.00; 1.00] |
| Park (2019) | 0.95 [0.92; 0.97] |
| Wellinghausen (2011) | 1.00 [0.99; 1.00] |
| Park (2019) | 0.83 [0.79; 0.86] |
| Total | 0.99 [0.98; 1.00] |
| Heterogeneity: $c^2_{14}$ = 396.12 ($P$ < .001), $I^2$ = 96% | |
| Total | 0.99 [0.98; 0.99] |

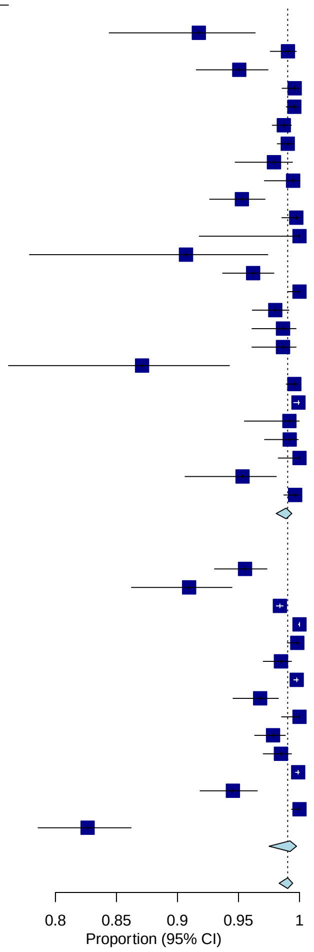

Heterogeneity: $c^2_{40}$ = 545.92 ($P$ < .001), $I^2$ = 93%
Test for subgroup differences: $c^2_1$ = 0.27 ($P$ = .60)

#### Data from the birth certificates

**Table S1: The number of detected syphilis cases in the birth certificate data.**

| Year | Race and Ethnicity | Number of detected syphilis infections | Reference |
|---|---|---|---|
| 2016 | Non-Hispanic American Indian/Alaska Native | 50 | 3 |
| 2016 | Non-Hispanic Asian | 97 | 3 |
| 2016 | Non-Hispanic Black/African American | 1533 | 3 |
| 2016 | Hispanic/Latino | 839 | 3 |
| 2016 | Non-Hispanic Multiracial | 93 | 3 |
| 2016 | Non-Hispanic Native Hawaiian/Other Pacific Islander | 45 | 3 |
| 2016 | Non-Hispanic White | 756 | 3 |
| 2017 | Non-Hispanic American Indian/Alaska Native | 68 | 4 |
| 2017 | Non-Hispanic Asian | 94 | 4 |
| 2017 | Non-Hispanic Black/African American | 1701 | 4 |
| 2017 | Hispanic/Latino | 1013 | 4 |
| 2017 | Non-Hispanic Multiracial | 75 | 4 |
| 2017 | Non-Hispanic Native Hawaiian/Other Pacific Islander | 45 | 4 |
| 2017 | Non-Hispanic White | 876 | 4 |
| 2018 | Non-Hispanic American Indian/Alaska Native | 70 | 5 |
| 2018 | Non-Hispanic Asian | 97 | 5 |
| 2018 | Non-Hispanic Black/African American | 1809 | 5 |
| 2018 | Hispanic/Latino | 1200 | 5 |
| 2018 | Non-Hispanic Multiracial | 126 | 5 |
| 2018 | Non-Hispanic Native Hawaiian/Other Pacific Islander | 46 | 5 |
| 2018 | Non-Hispanic White | 1031 | 5 |
| 2019 | Non-Hispanic American Indian/Alaska Native | 109 | 6 |
| 2019 | Non-Hispanic Asian | 127 | 6 |
| 2019 | Non-Hispanic Black/African American | 2250 | 6 |
| 2019 | Hispanic/Latino | 1514 | 6 |
| 2019 | Non-Hispanic Multiracial | 162 | 6 |
| 2019 | Non-Hispanic Native Hawaiian/Other Pacific Islander | 54 | 6 |
| 2019 | Non-Hispanic White | 1369 | 6 |
| 2020 | Non-Hispanic American Indian/Alaska Native | 150 | 7 |
| 2020 | Non-Hispanic Asian | 139 | 7 |
| 2020 | Non-Hispanic Black/African American | 2401 | 7 |
| 2020 | Hispanic/Latino | 1799 | 7 |
| 2020 | Non-Hispanic Multiracial | 192 | 7 |
| 2020 | Non-Hispanic Native Hawaiian/Other Pacific Islander | 61 | 7 |
| 2020 | Non-Hispanic White | 1662 | 7 |
| 2021 | Non-Hispanic American Indian/Alaska Native | 255 | 8 |
| 2021 | Non-Hispanic Asian | 131 | 8 |

| | | | |
|---|---|---|---|
| 2021 | Non-Hispanic Black/African American | 2824 | 8 |
| 2021 | Hispanic/Latino | 2073 | 8 |
| 2021 | Non-Hispanic Multiracial | 254 | 8 |
| 2021 | Non-Hispanic Native Hawaiian/Other Pacific Islander | 65 | 8 |
| 2021 | Non-Hispanic White | 2063 | 8 |
| 2022 | Non-Hispanic American Indian/Alaska Native | 361 | 9 |
| 2022 | Non-Hispanic Asian | 160 | 9 |
| 2022 | Non-Hispanic Black/African American | 3481 | 9 |
| 2022 | Hispanic/Latino | 2941 | 9 |
| 2022 | Non-Hispanic Multiracial | 311 | 9 |
| 2022 | Non-Hispanic Native Hawaiian/Other Pacific Islander | 72 | 9 |
| 2022 | Non-Hispanic White | 2803 | 9 |
| 2023 | Non-Hispanic American Indian/Alaska Native | 502 | 10 |
| 2023 | Non-Hispanic Asian | 173 | 10 |
| 2023 | Non-Hispanic Black/African American | 3842 | 10 |
| 2023 | Hispanic/Latino | 3388 | 10 |
| 2023 | Non-Hispanic Multiracial | 385 | 10 |
| 2023 | Non-Hispanic Native Hawaiian/Other Pacific Islander | 94 | 10 |
| 2023 | Non-Hispanic White | 3111 | 10 |

**Table S2: The number of people who are pregnant with live births in the United States.**

| Year | Race and Ethnicity | Population | Reference |
|---|---|---|---|
| 2016 | Non-Hispanic American Indian/Alaska Native | 31465 | [3] |
| 2016 | Non-Hispanic Asian | 254870 | [3] |
| 2016 | Non-Hispanic Black/African American | 559235 | [3] |
| 2016 | Hispanic/Latino | 926461 | [3] |
| 2016 | Non-Hispanic Multiracial | 80954 | [3] |
| 2016 | Non-Hispanic Native Hawaiian/Other Pacific Islander | 9350 | [3] |
| 2016 | Non-Hispanic White | 2057311 | [3] |
| 2017 | Non-Hispanic American Indian/Alaska Native | 29967 | [4] |
| 2017 | Non-Hispanic Asian | 249616 | [4] |
| 2017 | Non-Hispanic Black/African American | 561297 | [4] |
| 2017 | Hispanic/Latino | 905998 | [4] |
| 2017 | Non-Hispanic Multiracial | 82455 | [4] |
| 2017 | Non-Hispanic Native Hawaiian/Other Pacific Islander | 9436 | [4] |
| 2017 | Non-Hispanic White | 1993312 | [4] |
| 2018 | Non-Hispanic American Indian/Alaska Native | 29115 | [5] |
| 2018 | Non-Hispanic Asian | 241223 | [5] |
| 2018 | Non-Hispanic Black/African American | 552630 | [5] |
| 2018 | Hispanic/Latino | 893897 | [5] |
| 2018 | Non-Hispanic Multiracial | 83889 | [5] |
| 2018 | Non-Hispanic Native Hawaiian/Other Pacific Islander | 9481 | [5] |
| 2018 | Non-Hispanic White | 1957261 | [5] |

| | | | |
|---|---|---|---|
| 2019 | Non-Hispanic American Indian/Alaska Native | 28493 | 6 |
| 2019 | Non-Hispanic Asian | 239189 | 6 |
| 2019 | Non-Hispanic Black/African American | 548719 | 6 |
| 2019 | Hispanic/Latino | 893901 | 6 |
| 2019 | Non-Hispanic Multiracial | 84358 | 6 |
| 2019 | Non-Hispanic Native Hawaiian/Other Pacific Islander | 9778 | 6 |
| 2019 | Non-Hispanic White | 1917034 | 6 |
| 2020 | Non-Hispanic American Indian/Alaska Native | 26822 | 7 |
| 2020 | Non-Hispanic Asian | 219309 | 7 |
| 2020 | Non-Hispanic Black/African American | 530036 | 7 |
| 2020 | Hispanic/Latino | 871385 | 7 |
| 2020 | Non-Hispanic Multiracial | 84266 | 7 |
| 2020 | Non-Hispanic Native Hawaiian/Other Pacific Islander | 9630 | 7 |
| 2020 | Non-Hispanic White | 1844161 | 7 |
| 2021 | Non-Hispanic American Indian/Alaska Native | 26129 | 8 |
| 2021 | Non-Hispanic Asian | 213947 | 8 |
| 2021 | Non-Hispanic Black/African American | 518108 | 8 |
| 2021 | Hispanic/Latino | 890278 | 8 |
| 2021 | Non-Hispanic Multiracial | 87013 | 8 |
| 2021 | Non-Hispanic Native Hawaiian/Other Pacific Islander | 9536 | 8 |
| 2021 | Non-Hispanic White | 1888274 | 8 |
| 2022 | Non-Hispanic American Indian/Alaska Native | 25732 | 9 |
| 2022 | Non-Hispanic Asian | 219192 | 9 |

| | | | |
|---|---|---|---|
| 2022 | Non-Hispanic Black/African American | 511695 | 9 |
| 2022 | Hispanic/Latino | 944199 | 9 |
| 2022 | Non-Hispanic Multiracial | 88408 | 9 |
| 2022 | Non-Hispanic Native Hawaiian/Other Pacific Islander | 10128 | 9 |
| 2022 | Non-Hispanic White | 1841422 | 9 |
| 2023 | Non-Hispanic American Indian/Alaska Native | 24579 | 10 |
| 2023 | Non-Hispanic Asian | 216006 | 10 |
| 2023 | Non-Hispanic Black/African American | 491753 | 10 |
| 2023 | Hispanic/Latino | 952535 | 10 |
| 2023 | Non-Hispanic Multiracial | 89019 | 10 |
| 2023 | Non-Hispanic Native Hawaiian/Other Pacific Islander | 10124 | 10 |
| 2023 | Non-Hispanic White | 1787833 | 10 |

**Syphilis diagnoses in surveillance data.**

The diagnoses of primary, secondary, early, non-primary, and non-secondary syphilis among general women of reproductive ages (15-44 years) were derived from AtlasPlus database.[11] It served as the benchmark for the estimated prevalence of all stages of syphilis because the diagnoses for the late stage of syphilis and other stages of syphilis were not reported by the CDC.

**Table S3. Syphilis diagnoses among women of reproductive ages.**

| Year | Race and ethnicity | Diagnoses per 100,000 women of reproductive ages | Reference |
|---|---|---|---|
| 2016 | Non-Hispanic American Indian/Alaska Native | 7.7 | 11 |
| 2016 | Non-Hispanic Asian | 0.9 | 11 |
| 2016 | Non-Hispanic Black/African American | 15.2 | 11 |
| 2016 | Hispanic/Latino | 4.8 | 11 |
| 2016 | Non-Hispanic Multiracial | 2.8 | 11 |
| 2016 | Non-Hispanic Native Hawaiian/Other Pacific Islander | 12.2 | 11 |
| 2016 | Non-Hispanic White | 2.4 | 11 |
| 2017 | Non-Hispanic American Indian/Alaska Native | 16.7 | 11 |
| 2017 | Non-Hispanic Asian | 0.9 | 11 |
| 2017 | Non-Hispanic Black/African American | 16 | 11 |
| 2017 | Hispanic/Latino | 6 | 11 |
| 2017 | Non-Hispanic Multiracial | 3.3 | 11 |
| 2017 | Non-Hispanic Native Hawaiian/Other Pacific Islander | 10.4 | 11 |
| 2017 | Non-Hispanic White | 3.2 | 11 |
| 2018 | Non-Hispanic American Indian/Alaska Native | 21.5 | 11 |
| 2018 | Non-Hispanic Asian | 1.1 | 11 |
| 2018 | Non-Hispanic Black/African American | 20.1 | 11 |
| 2018 | Hispanic/Latino | 7.7 | 11 |
| 2018 | Non-Hispanic Multiracial | 5.6 | 11 |
| 2018 | Non-Hispanic Native Hawaiian/Other Pacific Islander | 14.7 | 11 |
| 2018 | Non-Hispanic White | 4.3 | 11 |
| 2019 | Non-Hispanic American Indian/Alaska Native | 35.6 | 11 |
| 2019 | Non-Hispanic Asian | 1.3 | 11 |
| 2019 | Non-Hispanic Black/African American | 23.9 | 11 |
| 2019 | Hispanic/Latino | 9.1 | 11 |
| 2019 | Non-Hispanic Multiracial | 7.4 | 11 |
| 2019 | Non-Hispanic Native Hawaiian/Other Pacific Islander | 15.2 | 11 |

| | | | |
|---|---|---|---|
| 2019 | Non-Hispanic White | 5.4 | 11 |
| 2020 | Non-Hispanic American Indian/Alaska Native | 45.6 | 11 |
| 2020 | Non-Hispanic Asian | 1.3 | 11 |
| 2020 | Non-Hispanic Black/African American | 27.7 | 11 |
| 2020 | Hispanic/Latino | 9.3 | 11 |
| 2020 | Non-Hispanic Multiracial | 9.7 | 11 |
| 2020 | Non-Hispanic Native Hawaiian/Other Pacific Islander | 28 | 11 |
| 2020 | Non-Hispanic White | 6.3 | 11 |
| 2021 | Non-Hispanic American Indian/Alaska Native | 78.6 | 11 |
| 2021 | Non-Hispanic Asian | 1.6 | 11 |
| 2021 | Non-Hispanic Black/African American | 35.6 | 11 |
| 2021 | Hispanic/Latino | 13.7 | 11 |
| 2021 | Non-Hispanic Multiracial | 20.4 | 11 |
| 2021 | Non-Hispanic Native Hawaiian/Other Pacific Islander | 32.2 | 11 |
| 2021 | Non-Hispanic White | 9.7 | 11 |
| 2022 | Non-Hispanic American Indian/Alaska Native | 119.9 | 11 |
| 2022 | Non-Hispanic Asian | 1.9 | 11 |
| 2022 | Non-Hispanic Black/African American | 39.7 | 11 |
| 2022 | Hispanic/Latino | 15.9 | 11 |
| 2022 | Non-Hispanic Multiracial | 27.8 | 11 |
| 2022 | Non-Hispanic Native Hawaiian/Other Pacific Islander | 31.7 | 11 |
| 2022 | Non-Hispanic White | 11.7 | 11 |
| 2023 | Non-Hispanic American Indian/Alaska Native | 110.6 | 11 |
| 2023 | Non-Hispanic Asian | 1.8 | 11 |
| 2023 | Non-Hispanic Black/African American | 39.8 | 11 |
| 2023 | Hispanic/Latino | 15.2 | 11 |
| 2023 | Non-Hispanic Multiracial | 32.3 | 11 |
| 2023 | Non-Hispanic Native Hawaiian/Other Pacific Islander | 35 | 11 |
| 2023 | Non-Hispanic White | 11.1 | 11 |

The diagnoses of all stages of syphilis among people who are pregnant were obtained from the Sexually Transmitted Surveillance 2022,[12] and 2023.[13]We compared our estimation against these values for all available years.

**Table S4. Syphilis diagnoses among people who are pregnant.**

| Year | Diagnoses per 100,000 | Reference |
|---|---|---|
| 2016 | 63.6 | 12,13 |
| 2017 | 84.2 | 12,13 |

| | | |
|---|---|---|
| 2018 | 100.0 | [12,13] |
| 2019 | 132.3 | [12,13] |
| 2020 | 158.8 | [12,13] |
| 2021 | 213.8 | [12,13] |
| 2022 | 268.4 | [12,13] |
| 2023 | 281.9 | [12,13] |

**Alternative weakly informative prior distributions for the syphilis prevalence among people who are pregnant for 2016–2023.**

**Table S5. Parameters used.**

| Parameter | Mean | Range | Prior distribution |
|---|---|---|---|
| Syphilis prevalence among people who are pregnant | 0.16 | 0.01–0.60 | Beta(1, 4) |
| Syphilis prevalence among people who are pregnant | 0.21 | 0.01–0.71 | Beta(1, 3) |
| Syphilis prevalence among people who are pregnant | 0.29 | 0.01–0.84 | Beta(1, 2) |
| Syphilis prevalence among people who are pregnant | 0.50 | 0.03–0.98 | Beta(1, 1) |

**The temporal change and racial/ethnic disparities in the estimated syphilis prevalence.**

**Figure S3. The temporal change in the estimated syphilis prevalence within each race/ethnicity group compared with 2014. The y-axis varies by subplot.**

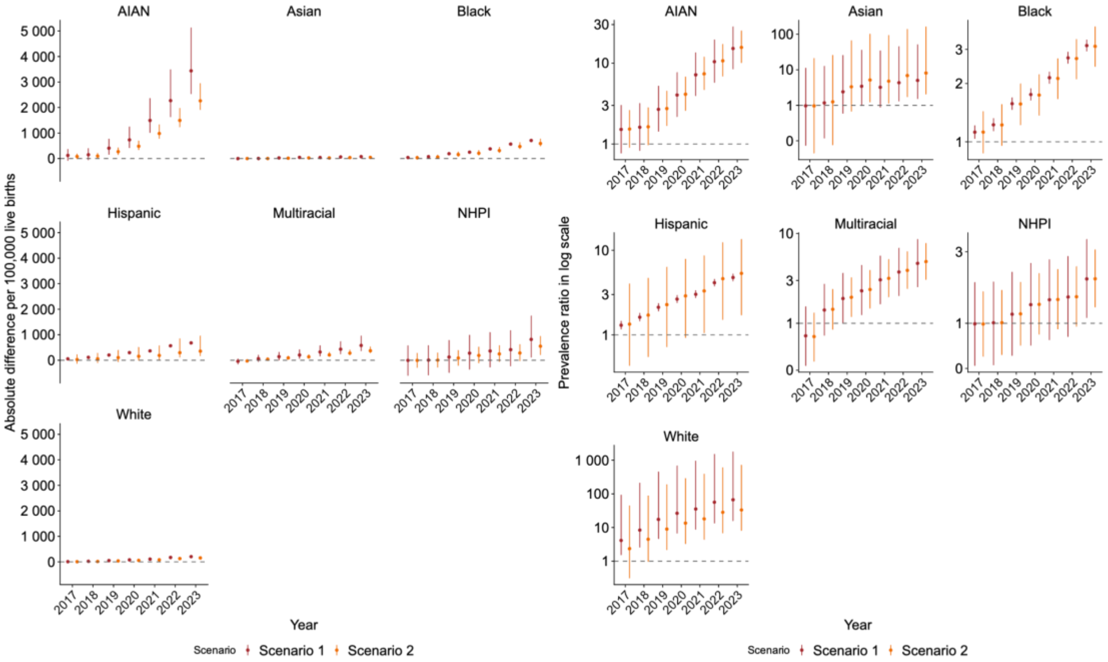

Footnote: In Scenario 1, we assume a lower screening coverage distribution among Medicare beneficiaries of each race/ethnicity. In Scenario 2, we assume a higher screening coverage distribution, ranging from the coverage reported by Medicare to the proportion of pregnant women receiving at least one prenatal care. Race and ethnicity categories: non-Hispanic American Indian/Alaska Native (AIAN), non-Hispanic Asian (Asian), non-Hispanic Black/African American (Black), Hispanic/Latino (Hispanic), non-Hispanic Multiracial (Multiracial), non-Hispanic Native Hawaiian/Other Pacific Islander (NHPI), and non-Hispanic White (White).

**Figure S4. Racial and ethnic disparities in the estimated syphilis prevalence compared with non-Hispanic white counterparts within the same year.**

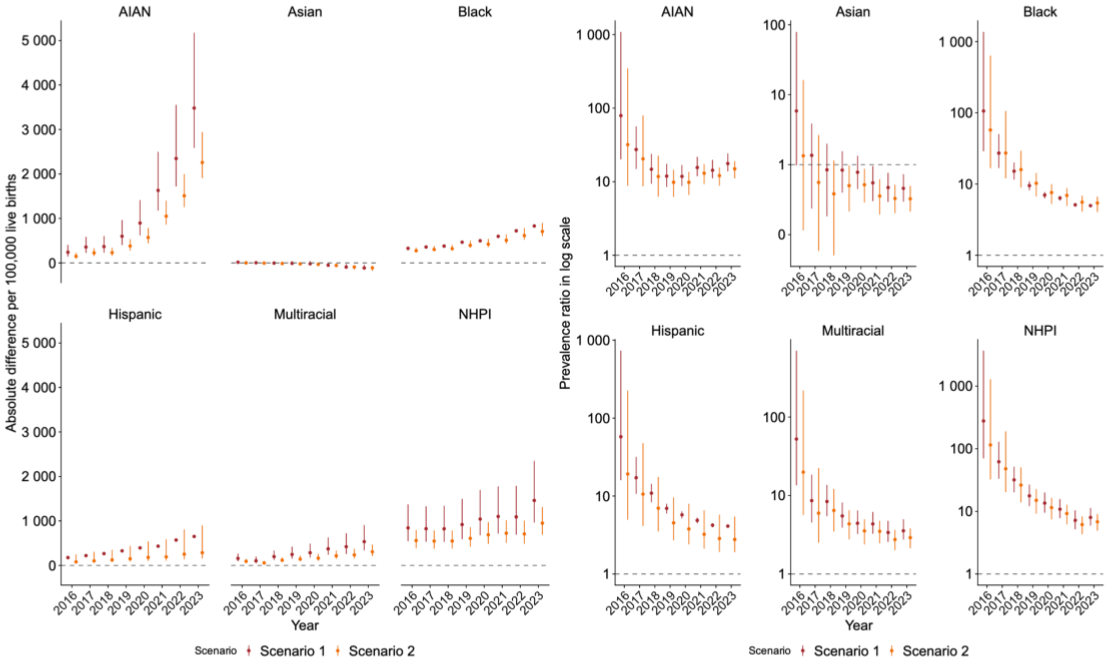

Footnote: In Scenario 1, we assume a lower screening coverage distribution among Medicare beneficiaries of each race/ethnicity. In Scenario 2, we assume a higher screening coverage distribution, ranging from the coverage reported by Medicare to the proportion of pregnant women receiving at least one prenatal care. Race and ethnicity categories: non-Hispanic American Indian/Alaska Native (AIAN), non-Hispanic Asian (Asian), non-Hispanic Black/African American (Black), Hispanic/Latino (Hispanic), non-Hispanic Multiracial (Multiracial), non-Hispanic Native Hawaiian/Other Pacific Islander (NHPI), and non-Hispanic White (White).

**The estimated syphilis prevalence per 100,000 live births by race and ethnicity with different weekly informative prior distributions for 2016–2023.**

**Figure S5. Sensitivity analysis: the estimated syphilis prevalence per 100,000 live births with alternative weakly informative prior distributions for the syphilis prevalence for 2016–2023. The y-axis varies by subplot. (Top: Scenario 1; Bottom: Scenario 2).**

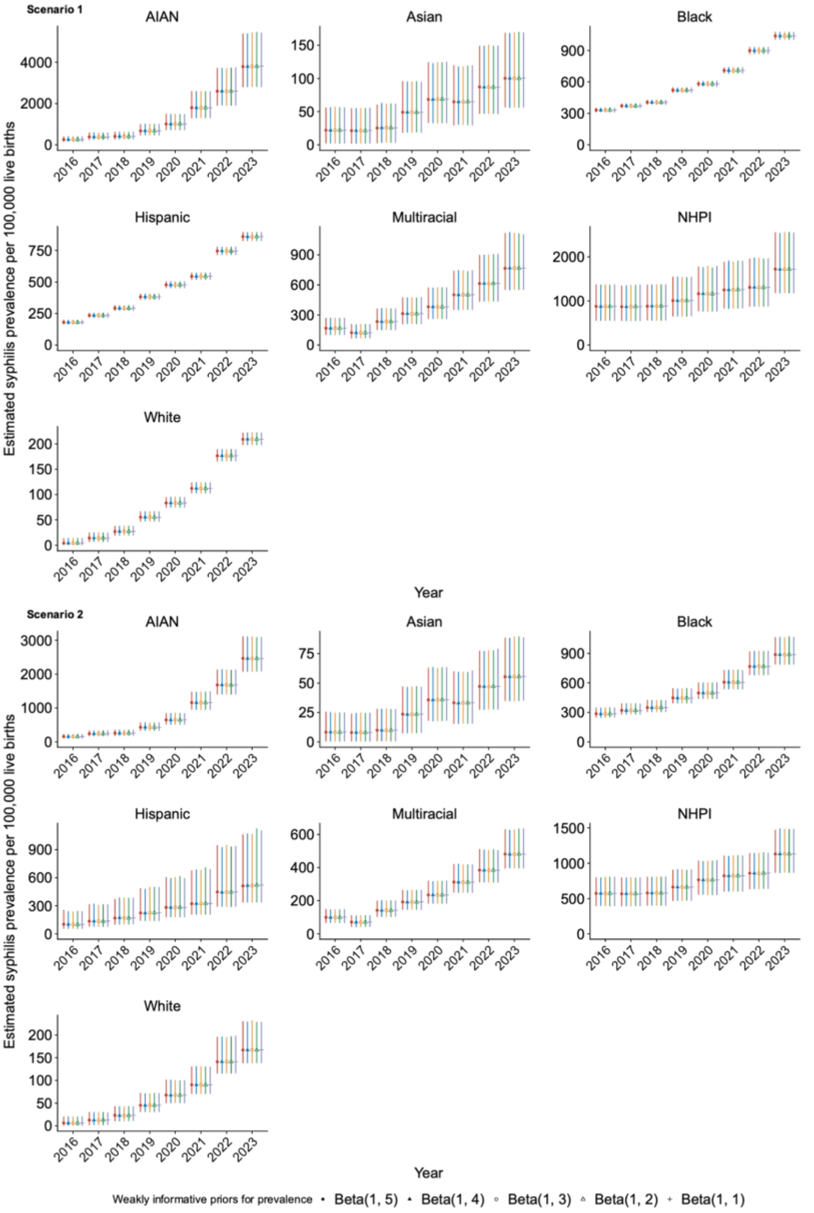

Footnote: In Scenario 1, we assume a lower screening coverage distribution among Medicare beneficiaries of each race/ethnicity. In Scenario 2, we assume a higher screening coverage distribution, ranging from the coverage reported by Medicare to the proportion of pregnant women receiving at least one prenatal care. Race and ethnicity categories: non-Hispanic American Indian/Alaska Native (AIAN), non-Hispanic Asian (Asian), non-Hispanic Black/African American (Black), Hispanic/Latino (Hispanic), non-Hispanic Multiracial (Multiracial), non-Hispanic Native Hawaiian/Other Pacific Islander (NHPI), and non-Hispanic White (White).
